# Comparing Adverse Maternal And Neonatal Outcomes in Adolescent versus Adult Mothers in Sub-Saharan Africa

**DOI:** 10.1101/2024.01.04.24300852

**Authors:** Chibuikem N. Onuzo, Promise E. Sefogah, Nelson K. Damale, Theodore K. Boafor, Alim Swarray-Deen, Kareem Mumuni

## Abstract

**Objective:** Adolescent pregnancy is a major social and public health problem that burdens affected families, the communities and societies globally. It has been associated with a higher prevalence of adverse pregnancy outcomes compared to pregnancy in adults. To compare adverse pregnancy outcomes in adolescents (13-19 years) and those in adults (20 to 35 years) at the Korle-Bu Teaching Hospital in Accra, Ghana and investigate the associated factors among adolescents.

**Methods:** This comparative cross-sectional study involved 110 adolescents (13-19 years) and 220 adults (20 to 35 years) who delivered at the Maternity Unit of the Korle-Bu Teaching Hospital between November 2016 and February 2017. Participants were recruited using the convenience sampling method. After study protocol was explained to the parturient, informed voluntary consent and assent were obtained. Participants who met the inclusion criteria were recruited in the study. Sociodemographic characteristics, antenatal and delivery records; and maternal and perinatal outcomes were collected using an interviewer administered questionnaire and the participants’ records. Data analysis was done using SPSS version 16.0.

**Results:** The incidence of adolescent pregnancies during the study period was 5.1%. Compared to adults, adolescents were about 3 times more likely to have eclampsia although preeclampsia occurred more in adults. Adolescents that resided in sub-urban dwellings were more likely to have an adverse perinatal outcome compared to their adult counterparts.

**Conclusion:** Our study found that, in addition to their socioeconomic and biophysical disadvantages, adolescents were likely to have exceptionally high risk of eclampsia.

## Introduction

Adolescence is a critical transitional period in life characterized by a tremendous pace in growth and body change that is second only to that of infancy [1]. Worldwide, about 16 million girls aged 15 to 19 years, and 1 million girls under 15 years give birth every year, accounting for 11% of total births worldwide with 95% of these births occurring in low– and middle-income countries [1][2]. Adolescent pregnancy poses significantly high risk to both mother and baby including cephalopelvic disproportion, increased risk of caesarean sections, hypertensive disorders (including eclampsia) and maternal mortality. Others include perinatal complications such as low birth weight, preterm birth and perinatal mortality [1][2][3]. The reasons for association between a younger maternal age and these adverse pregnancy outcomes remain controversial as some evidence points to the socioeconomic disadvantage while others suggest biological immaturity [2]. Poverty, low literacy levels, unemployment and sub-urban residence are major predisposing factors to adolescent pregnancy [1][2]. Others risk factors include living with single parents, inadequate family support and child marriage [2][3].

## Objective

To compare adverse pregnancy outcomes between adolescents (13 to 19 years) and adults (20 to 34 years) at the largest teaching hospital in Ghana and investigate the factors associated with these outcomes among adolescents.

## Methodology

The study was a Comparative Cross-Sectional study involving two groups of pregnant women; adolescents aged 13 to 19 years and adults 20 to 35years old. The study was conducted at the Maternity Unit of the Department of Obstetrics and Gynaecology, Korle-Bu Teaching hospital over a period of 14 weeks (November 11, 2016 – February 16, 2017). The study was approved by the Ethical and Protocol Review Committee of the College of Health Sciences, Korle Bu. (ERC-Et/M.8-P4.3/2015-2016)

The sample size for the adolescent group was calculated using the formula N = Z^2^ x (Pq) / E^2^. The total sample size of 330 was calculated for the study comprising 110 adolescents and 220 adults using a 1:2 ratio. Adolescents who delivered over each preceding 24-hour period were identified and the next two adults in the register who met the inclusion criteria were selected into the comparative group by the convenience sampling method. Written consent was given by all adolescents over 18 years and adults in the study. Parents (or in-loco-parentis) of adolescents less than 18years also gave consent for their wards while the adolescents assented to participation in the study. Participants’ sociodemographic data were obtained from their clinical records followed by administration of structured pre-tested questionnaires administered to each participant. The clients and their babies were retained in the study for up for one week following delivery and any adverse developments reported. Data was entered in Microsoft excel and exported into SPSS version 16 for analysis. Categorical variables were compared using chi-squared and Fisher’s exact tests while the numerical variables were compared using the students t-test and the Mann-Witney U tests.

Odds ratio (OR) with 95% confidence intervals were calculated to measure the association between variables and the maternal age. Logistic regression analysis was performed to determine the predictors of the significant outcomes in both groups and a composite of adverse maternal and neonatal outcomes in the adolescent group. Results were considered statistically significant if the p-value was less than 0.05.

## Results

Adolescent deliveries constituted (110) 5.1% of the study participants and there were 4 (0.18%) early adolescents (aged 15-years and below), 3.6% of the adolescent group. The mean age of the adolescents was 17.6 (SD 1.2) years, whiles that of the adults was 30.4 (SD 5.0) years. Compared to adults, significantly more adolescents were unmarried (17.7% vs 86.4%), cohabiting (5.9% vs 10.9%), had unplanned pregnancies, were suburban residents (7.3% vs 24.5%) students (1.9% vs 33.3%), did not attain tertiary education (34.5% vs 3.6%) and were unemployed (1.9% vs 26.7%). (Table 1)

**Table 1:** Socio-demographic characteristics.

| **Variable** | **Adults**<br>**N=220**<br>**n(%) / mean±SD** | **Adolescents**<br>**N=110**<br>**n(%) / mean±SD** | **Total**<br>**N=330**<br>**n(%) / mean±SD** | **X-square/**<br>**Fisher's**<br>**Value** | **P-**<br>**value** |
| --- | --- | --- | --- | --- | --- |
| **Mean Age** | 30.4 ±5.0 | 17.6 ±1.2 | 26.1 ±7.3 | - | **<0.00#** |
| **Marital status** |  |  |  | **150.49** | **<0.001** |
| Married | 181 (82.3) | 15 (13.6) | 196 (59.4) |  |  |
| Single | 26 (11.8) | 83 (75.5) | 109 (33.0) |  |  |
| Co-habiting | 13 (5.9) | 12 (10.9) | 25 (7.6) |  |  |
| **Highest level of**<br>**education** |  |  |  | **41.661** | **<0.001** |
| No formal<br>education | 20 (9.1) | 8 (7.3) | 28 (8.6) |  |  |
| Primary | 21 (9.5) | 21 (19.1) | 42 (12.7) |  |  |
| Secondary | 103 (46.8) | 77 (70.0) | 180 (54.5) |  |  |
| Tertiary | 76 (34.5) | 4 (3.6) | 80 (24.2) |  |  |
| **Occupation** | **N=216** | **N=105** | **N=321** | **140.70** | **<0.001** |
| Trader/Artisan | 138 (63.8) | 41 (39.0) | 179 (55.7) |  |  |
| Professional | 70 (32.4) | 1 (1.0) | 71 (22.1) |  |  |
| Student | 4 (1.9) | 35 (33.3) | 39 (12.1) |  |  |
| Unemployed | 4 (1.9) | 28 (26.7) | 32 (10.0) |  |  |
| **Residence** |  |  |  | **19.306** | **<0.001** |
| Suburban | 16 (7.3) | 27 (24.5) | 43 (13.0) |  |  |
| Urban | 204 (92.7) | 83 (75.5) | 287 (87.0) |  |  |

Most adolescents had their first sexual experience at a relatively younger age 16.3 (± 1.4 years) compared to adults 21.1 (± 2.6 years) but were less likely to have had two or more lifetime sexual partners. Adolescents booked later and attended significantly fewer antenatal clinic reviews than the adults. Contraceptive knowledge and uptake was significantly lower among adolescents and they also had a lower mean Haemoglobin levels at booking (10.0g/dL ±1.6g/dL) compared to adults (10.6g/dL ± 1.3g/dL). Although preeclampsia occurred more in adults than adolescents (7.7% vs 6.4%), eclampsia was the only adverse maternal outcome found to be significantly higher among adolescents, occurring about three times more in the adolescents [OR = 3.37, 95% C.I. (1.08 – 10.57)] compared to adults. There was no significant difference in the proportions of peripartum complications including cephalo-pelvic disproportion (CPD), antepartum haemorrhage (APH), postpartum haemorrhage (PPH) and premature rupture of membranes (PROM) between the two groups. (Table 2), (Table 3)

**Table 2:** Maternal Outcomes (labour and delivery)

| **Variable** | **Adults**<br>**N=220**<br>**n (%)** | **Adolescents**<br>**N=110**<br>**n (%)** | **OR (95% C.I)** | **X<sup>2</sup>/**<br>**Fisher's**<br>**Value** | **P-**<br>**value** |
| --- | --- | --- | --- | --- | --- |
| **Patients that laboured** |  |  | 1.66 (0.87-3.20) | 1.933 | 0.164 |
| Yes | 177 (80.5) | 96 (87.3) |  |  |  |
| No | 43 (19.5) | 14 (12.7) |  |  |  |
| **Onset of labour** | **N=177** | **N=96** | 0.88 (0.37-2.09) | 0.257 | 0.669 |
| Spontaneous | 155 (87.6) | 86 (89.5) |  |  |  |
| Induced | 22 (12.4) | 10 (11.5) |  |  |  |
| **Labour augmentation** | **N=177** | **N=96** | 1.43 (0.73-2.77) | 0.767 | 0.381 |
| No | 147 (83.1) | 76 (79.2) |  |  |  |
| Yes | 30 (16.9) | 20 (20.8) |  |  |  |
| **Outcome of labour** |  |  |  |  |  |
| Successful vaginal birth | 147 (83.1) | 80 (83.3) | 1.02 (0.52 – 1.98) | 0.003 | 0.952 |
| Emergency CS | 30 (16.9) | 16 (16.7) |  |  |  |
| **Mode of delivery** |  |  | 1.34 (0.80-2.19) | 1.708 | 0.334 |
| Caesarean section | 73 (33.2) | 30 (27.3) |  |  |  |
| Vaginal delivery | 147 (66.8) | 80 (72.7) |  |  |  |
| **Type of CS** | **N=73** | **N=30** | 1.97 (0.60-6.45) | 1.298 | 0.296* |
| Emergency | 56 (76.7) | 26 (86.7) |  |  |  |
| Elective | 17 (23.3) | 4 (13.3) |  |  |  |
| **Emergency CS indication** | **N = 56** | **N = 26** |  |  |  |
| Complications of labour | 30 | 16 |  |  |  |
| Other indications | 26 | 10 |  |  |  |

**Table 3:**
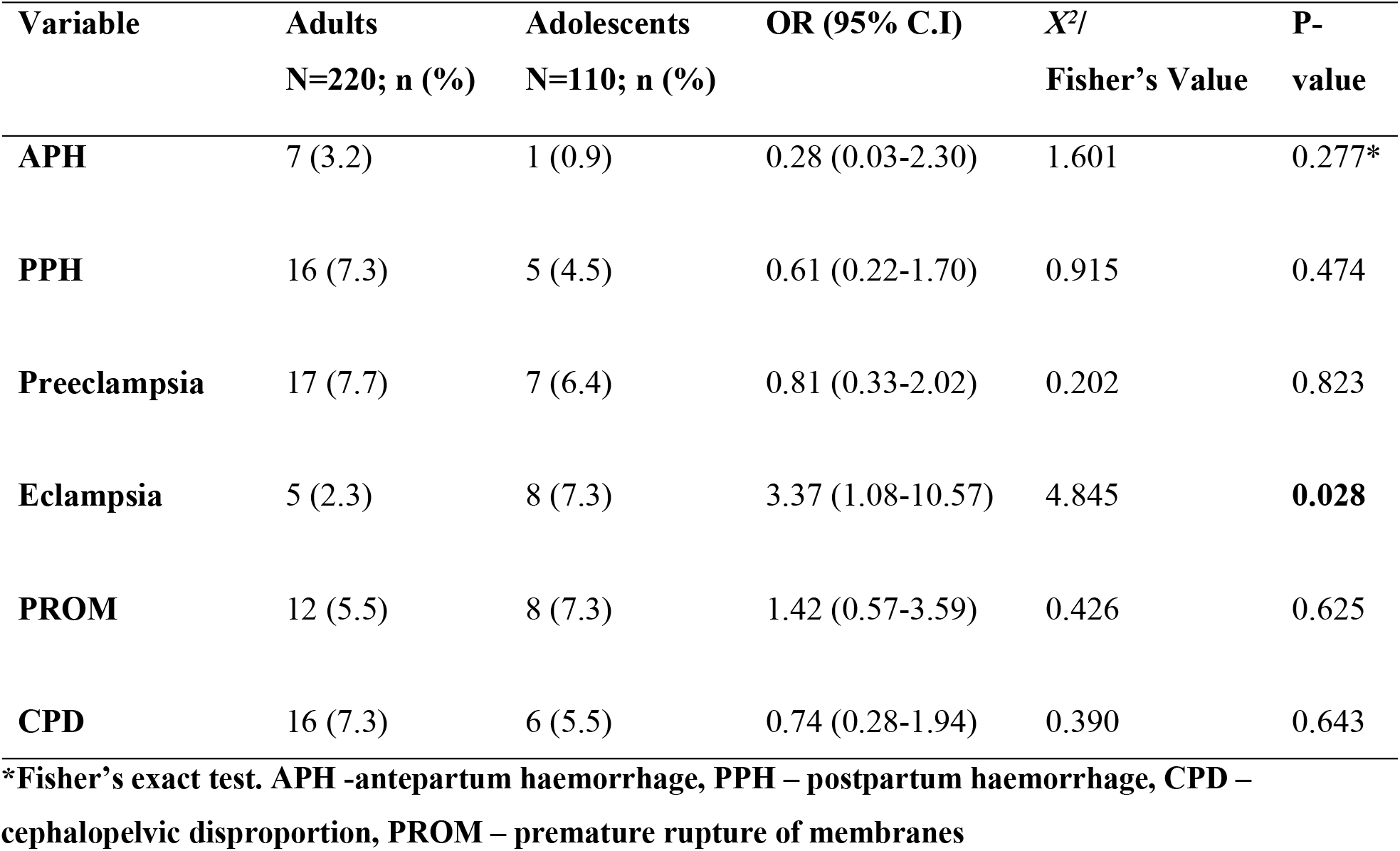
Maternal Outcomes (Antenatal)

The onset of labour, need for labour induction and augmentation, mode of delivery and successful vaginal births were similar for both groups of mothers. There was also no significant difference in the prevalence of syphilis or HIV infection between both groups although hepatitis B virus infection (HBV) prevalence was significantly higher among adults (10.5%) compared to adolescents (1.8%). (p<0.004) (Table 4)

**Table 4:** Maternal Infections. * *Fisher’s exact test.* HIV – Human Immunodeficiency Virus

| Variable | Adults<br>N=220; n(%) | Adolescents<br>N=110; n (%) | OR (95% C.I) | X<sup>2</sup> /<br>Fisher's<br>Value | P-value |
| --- | --- | --- | --- | --- | --- |
| **HIV infection** | 3 (1.4) | 2 (1.8) | 1.34 (0.22-8.14) | 0.102 | 1.000 |
| **Syphilis** | 4 (1.8) | 4 (3.6) | 2.04 (0.50-8.31) | 1.025 | 0.448* |
| **Hepatitis B infection** | 23 (10.5) | 2 (1.8) | 0.16 (0.04-0.69) | **7.812** | **0.004\*** |
| **Urinary tract infection** | 49 (22.3) | 13 (11.8) | 0.47 (0.24-0.91) | **5.253** | **0.025** |

Perinatal Outcomes (Birth Weight, Preterm Births, Stillbirths)

There was no significant difference in the mean birth weight of babies born to adolescents (2.7kg ± 0.7kg) and adults (2.9kg ± 0.7kg). (Table 5)

**Table 5:** Perinatal Outcomes (birth weight, preterm birth, still births)

| **Variable** | **Adults**<br>**N=220**<br>**n(%) / mean±SD** | **Adolescents**<br>**N=110**<br>**n(%) / mean±SD** | **OR**<br>**(95% C.I)** | **X<sup>2</sup>/**<br>**Fisher's** | **P-value** |
| --- | --- | --- | --- | --- | --- |
| **Mean Birth weight** | 2.9 ±0.7 | 2.7 ±0.7 | 2.8 ±0.7 | 1.583 | 0.114 |
| **Birth weight class** |  |  | 0.88 (0.52-1.51) | 0.201 | 0.654 |
| **LBW (<2.5kg)** | 57 (25.9) | 26 (23.6) |  |  |  |
| **NBW (>2.5kg)** | 163 (74.1) | 84 (76.4) |  |  |  |
| **Preterm birth** |  |  | 0.81 (0.39-1.65) | 0.348 | 0.600* |
| No | 191 (86.8) | 98 (89.1) |  |  |  |
| Yes | 29 (13.2) | 12 (10.9) |  |  |  |
| **Preterm class** | **N=29** | **N=12** | 0.67 (0.17-2.60) | 0.344 | 0.734 |
| 28- 32 weeks | 15 (51.7) | 5 (41.7) |  |  |  |
| 32- 36 weeks | 14 (48.3) | 7 (58.3) |  |  |  |
| **Still births** |  |  | 1 (0.37-2.74) |  |  |
| No | 208 (94.5) | 104 (94.5) |  | 0.000 | 1.000 |
| Yes | 12 (5.5) | 6 (5.5) |  |  |  |
| **Fresh still births** |  |  | 1.70 (0.51-5.69) |  |  |
| No | 214 (97.3) | 105 (95.5) |  | 0.294 | 0.516 |
| Yes | 6 (2.7) | 5 (4.5) |  |  |  |
| **Stillbirth class** | **N=12** | **N=6** | 5.00 (0.44-56.63) | 0.731 | 0.316* |
| **FSB** | 6 (50.0) | 5 (83.3) |  |  |  |
| **MSB** | 6 (50.0) | 1 (16.7) |  |  |  |

*\*Fisher's exact test, #t-test. Tests are significant at p<0.05. BW – Birth Weight, LBW – Low birth weight, NBW – Normal birth weight, FSB – Fresh stillbirth, MSB – Macerated stillbirth.*

There was no significant difference in the rates of preterm births, low birth weight and still birth between the two groups. There were also no significant differences in the proportions of babies born with low APGAR scores, babies admitted to the neonatal intensive care unit (NICU) and indications for NICU admissions. (Table 6)

**Table 6:** Perinatal Outcomes (Apgar, NICU admission, perinatal mortality) * *Fisher’s exact test*. Tests are significant at p<0.05

| Variable | Adults<br>N=220<br>n (%) | Adolescents<br>N=110<br>n (%) | OR (95% C.I) | X<sup>2</sup>/<br>Fisher's | P-value |
| --- | --- | --- | --- | --- | --- |
| **1 minute Apgar** | **N=199** | **N=104** | 0.86 (0.53-1.39) | 0.397 | 0.529 |
| <7 | 115 (57.8) | 64 (61.5) |  |  |  |
| ≥7 | 84 (42.2) | 40 (38.5) |  |  |  |
| **5 minute Apgar** | **N=199** | **N=104** | 0.68 (0.33-1.42) | 1.078 | 0.333 |
| <7 | 19 (9.5) | 14 (4.8) |  |  |  |
| ≥7 | 180 (90.5) | 90 (86.5) |  |  |  |
| **NICU admission** |  |  | 1.20 (0.63-2.30) | 0.959 | 0.351 |
| No | 191 (86.8) | 93 (84.5) |  |  |  |
| Yes | 29 (13.2) | 17 (15.5) |  |  |  |
| **Indication for**<br>**NICU admission** |  |  |  | 5.043 | 0.411* |
| Premature | 17 (51.5) | 7 (41.2) |  |  |  |
| Asphyxia/RDS | 8 (29.6) | 14 (47.1) |  |  |  |
| Macrosomia | 4 (14.8) | 0 (0.0) |  |  |  |
| Mother with DM | 4 (14.8) | 2 (11.8) |  |  |  |
| **Early neonatal**<br>**death** | 3 (1.4) | 3(2.7) | 2.03 (0.40-10.22) | 0.764 | 0.404 |
| **Perinatal deaths** | 15 (6.8) | 9 (8.2) | 0.82 (0.35-1.94) | 0.202 | 0.822 |

RDS – Respiratory distress syndrome, DM – Diabetes Mellitus, NICU – Neonatal intensive care unit.

There was no significant difference in the proportions of early neonatal deaths (p=0.404) or perinatal deaths (p=0.822) between babies of adolescents and those of adults. Of all the sociodemographic, medical and obstetric parameters analysed, only the diagnosis of preeclampsia was a significant predictor of eclampsia. (Table 7)

**Table 7:**
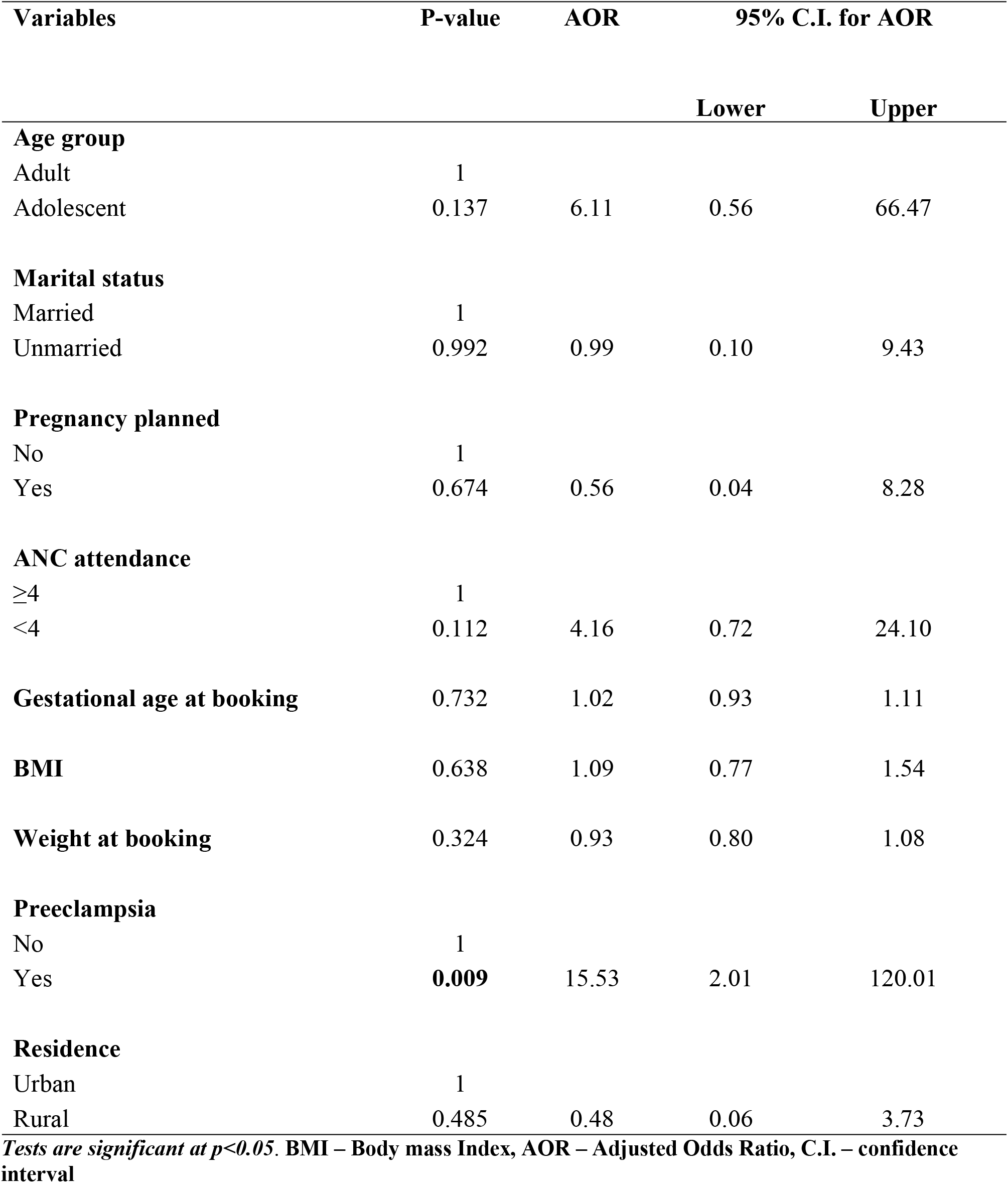
Logistic regression analysis of selected factors associated with eclampsia.

| Variables | P-value | AOR | 95% C.I. for AOR |  |
| --- | --- | --- | --- | --- |
|  |  |  | Lower | Upper |
| Age group |  |  |  |  |
| Adult | 1 |  |  |  |
| Adolescent | 0.137 | 6.11 | 0.56 | 66.47 |
| Marital status |  |  |  |  |
| Married | 1 |  |  |  |
| Unmarried | 0.992 | 0.99 | 0.10 | 9.43 |
| Pregnancy planned |  |  |  |  |
| No | 1 |  |  |  |
| Yes | 0.674 | 0.56 | 0.04 | 8.28 |
| ANC attendance |  |  |  |  |
| ≥4 | 1 |  |  |  |
| <4 | 0.112 | 4.16 | 0.72 | 24.10 |
| Gestational age at booking | 0.732 | 1.02 | 0.93 | 1.11 |
| BMI | 0.638 | 1.09 | 0.77 | 1.54 |
| Weight at booking | 0.324 | 0.93 | 0.80 | 1.08 |
| Preeclampsia |  |  |  |  |
| No | 1 |  |  |  |
| Yes | 0.009 | 15.53 | 2.01 | 120.01 |
| Residence |  |  |  |  |
| Urban | 1 |  |  |  |
| Rural | 0.485 | 0.48 | 0.06 | 3.73 |

*Tests are significant at $p < 0.05$ . BMI – Body mass Index, AOR – Adjusted Odds Ratio, C.I. – confidence interval*

## Discussion

Our study compared adverse maternal and neonatal outcomes among adolescent and adult parturient at the Korle Bu Teaching Hospital. Adolescents are considered biologically immature and psychologically unprepared to handle the physical and emotional demands of pregnancy. They are also socially disadvantaged, commonly occupy the lower end of the wealth quartile and lack the economic empowerment required for the demands of pregnancy. The prevalence of adolescent pregnancy from this study (5.1%) is similar to findings from a retrospective, hospital-based study in Southern Nigerian which reported a prevalence of 6.2% [4]. A number of other Teaching Hospital studies from Cameroon, Southwest and Southeast Nigeria reported lower adolescent pregnancy rates of 1.3%, 2.84%, 2.25% and 1.67% respectively [5][6][7][8]. Ogunlesi et al reported that adolescents in Sagamu were unlikely to visit the Teaching Hospital unless they were referred due to shame arising from cultural disapproval of teenage pregnancy [7].

Unfavourable socioeconomic circumstances of pregnant adolescents including being unmarried, unemployed, attaining lower levels of education and residing in suburban areas, as found in this study have been consistently reported as major predispositions to adverse pregnancy outcomes among adolescents [5][9][10]. Although a large proportion of adolescent pregnancies were unplanned, almost all the adolescents in this study made at least one visit to the antenatal clinic. The proportion of non-attendant adolescents in this study (2.7%) was significantly lower than those from similar studies in Sagamu (76%), Abakiliki (23.4%) Calabar (62.0%) and Kuala Lumpur (26.9%) [6][7][11][12]. This may be a reflection of the general improvement in antenatal coverage in Ghana from 82% in 1988 to 97% in 2014 and the free maternal healthcare policy under Ghana’s National Health Insurance Scheme (NHIS) [13].

Eclampsia was the only adverse pregnancy outcome found to be significantly higher (about five times) among adolescents compared to adults in this study. This agrees with findings by Ampofo et al at the same hospital in 1985 [14]. However, our current study found no significant difference in the incidence of preeclampsia between the two groups, although adolescents were about forty-times more likely to be nulliparous, a known risk factor for preeclampsia [2]. These findings may be explained by the large proportion (75%) of adolescents who were referred to the KBTH following an eclamptic fit. Similar findings were reported by Ezegwui, Conde-Agudelo and Gupta [5][15][16]. Poor or no antenatal attendance was the major predisposition to eclampsia among adolescents in Ampofo’s study [14]. They drew a correlation between unfavourable socioeconomic circumstances which prevailed in Ghana at the time of their study and the higher prevalence of eclampsia among adolescents as they tended to be poorer, less educated, unemployed and belonged to the lower end of the wealth quartile compared to their adult counterparts [14]. More than thirty years on, eclampsia is still being diagnosed in comparatively higher proportions among adolescents despite improvements in the general economic circumstances in Ghana. Seeing complications of eclampsia have been reported by Bonsaffoh et al as major contributors to maternal mortality at the KBTH, this high prevalence of eclampsia among these adolescents would imply greater inherent risk they face from dying through these complications [17]. This would further necessitate the call for primary prevention of pregnancy among adolescents in order to avert these avoidable maternal deaths.

## Conclusion

Our study found that, in addition to their socioeconomic and biological disadvantages, adolescents have exceptionally high risk of eclampsia. Other maternal and neonatal outcomes were similar to those in adults. Efforts at preventing adolescent pregnancy would hold significant prospects for reducing the burden of eclampsia and its complications including maternal mortality.

## Authors’ contributions

CNO, NKD and PES co-designed the study and questionnaire. CNO, PES and KM participated in proposal writing, data collection and analysis. CNO, ASD, NKD and KM, supervised proposal writing, data collection and writing protocol. CNO, PES and TKB performed and reviewed the data analysis. CNO, PES, TKB and ASD drafted the first version of the manuscript and PES is responsible for correspondence. All authors participated in manuscript writing, review and approved the final manuscript.

## Funding

This study received no funding.

## Conflict of Interest

Authors have no conflicts of interest to declare.

## Data Availability

All relevant data are within the manuscript and its Supporting Information files.

## Acknowledgements

Our sincere appreciations to the doctors, midwives and nurses at the obstetric unit of the Korle Bu Teaching Hospital for their immense support during this study.

## Notes

### Competing Interest Statement

The authors have declared no competing interest.

### Funding Statement

The author(s) received no specific funding for this work.

### Author Declarations

The study was approved by the Ethical and Protocol Review Committee of the College of Health Sciences, Korle Bu. (ERC-Et/M.8-P4.3/2015-2016)

